# Water fluoridation : A bibliometric study from 1950 to 2020

**DOI:** 10.1101/2023.11.07.23298219

**Authors:** Anqi Liu, Changyong Yuan, Ling Xu, Li Zhao

## Abstract

Water fluoridation (WF) is considered by the Centers for Disease Control and Prevention (CDC)to be one of the top ten public health achievements of the 20th century. However, discussions about WF have been ongoing. The purpose of this study is to collect and analyze the scientific literature for WF and understand the keyword network of the entire field, important articles, high-yield countries, institutions and authors, and the mutual cooperation between them. The data of this research are extracted from the Web of Science (WoS) from 1950 to 2020. In total, 1,008 articles were published. The United States published the largest number of papers. Australia’s University of Adelaide ranks first among the top 20 institutions. Seven scholars, including AJ Spencer, constitute the largest author collaboration network. Community Dentistry and Oral Epidemiology had the most publications, with 107 articles. Keywords density map found that the main keywords were dental caries, fluoride, children, drinking water, prevalence, and dental fluorosis. Keywords clusters mainly related to dental caries, fluoride, epidemiology research, and dental fluorosis. In the highly cited literature, we found that the research also focused on the relationship between stopping WF and caries and the systemic effects of WF.

## Introduction

In 2021, the U.S. Centers for Disease Control and Prevention (CDC) celebrated the 75th anniversary of water fluoridation (WF).^1^ By 2018, 73% of the U.S. population has drunk fluoridated water, and there are plans to increase this proportion to 80% by 2020.^2^ WF is considered by the CDC to be one of the top ten public health achievements of the 20th century.^3^ Numerous studies have reported the effectiveness of WF in caries prevention.^4,5^

However, discussions about WF have been ongoing, and both supporters and opponents in the academic community have put forward their own views.^6^ An investigation of public tweets on WF shows that the words “poisoning” and “waste” frequently appear, prompting the public to be strongly concerned about health damage that WF may cause.^7^ In view of the long-term widespread use of WF, there is controversy. It is necessary to conduct detailed bibliometric research on the entire WF literature, to understand the main status of current scientific research and to provide references for later research. Bibliometric analysis is widely used in various fields, including management,^8^ environmental sciences,^9^ medicine,^10^ etc. Scientific bibliometrics can involve statistical analysis on thousands to tens of thousands of documents. By measuring the key information among them, a mathematical matrix is formed and then visualized as a network diagram. By simplifying complex information, the main structure can be understood and analysed by scholars.

VOSviewer was designed by NJ van Eck and L Waltman of Leiden University in the Netherlands, specifically for the visual analysis of bibliometrics.^11^ Therefore, in this study, data were organized on Web of Science (WoS), and then VOSviewer was used for co-authorship analysis and keyword analysis. HistCite was designed by Garfield and used for document analysis and statistical analysis of highly cited literature.^12^

The purpose of this study is to collect and measure scientific literature data on WF and elucidate the keyword network of the entire field, important articles, high-yield countries, institutions and authors, and the mutual cooperation between them.

## Methods

### Data Source and Search Strategy

We chose the Science Citation Index Expanded (SCI-EXPANDED) from the WoS Core Collection (WoSCC). The search formula was as follows: TS=(“water fluoridation”) OR TS=(“water-fluoridation”) OR TS=(“fluoridated water”) OR TS=(“fluoridated-water”) OR TS=(“fluoride water”) OR TS=(“fluoride-water”) OR TS=(“water-fluoridation”) OR TS=(“fluoridated water”) OR TS=(“fluoridated-water”).

Topic research refers to searching for all topic-related parts, including the title, abstract and keywords. The time span of the selected literature is from 1950 to 2020.

### Data Collection

After obtaining the preliminary search results, we further refined them. The refinement process involved selecting “articles” and “reviews” through the document type and selecting “English” as the language of the article. Our search time was January 21, 2021. Two experts independently determined whether the document should be excluded by browsing the title and abstract of the document. When there was a discrepancy between the two experts, through discussion, it was determined whether the document was finally excluded. “Full Record and Cited References” were selected in the retrieval data export, and “Plain Text” was selected as the data format for VOSviewer and HistCite. Figure 1 shows the flow of the retrieval strategy and the structure diagram of this article.

**Figure 1.**
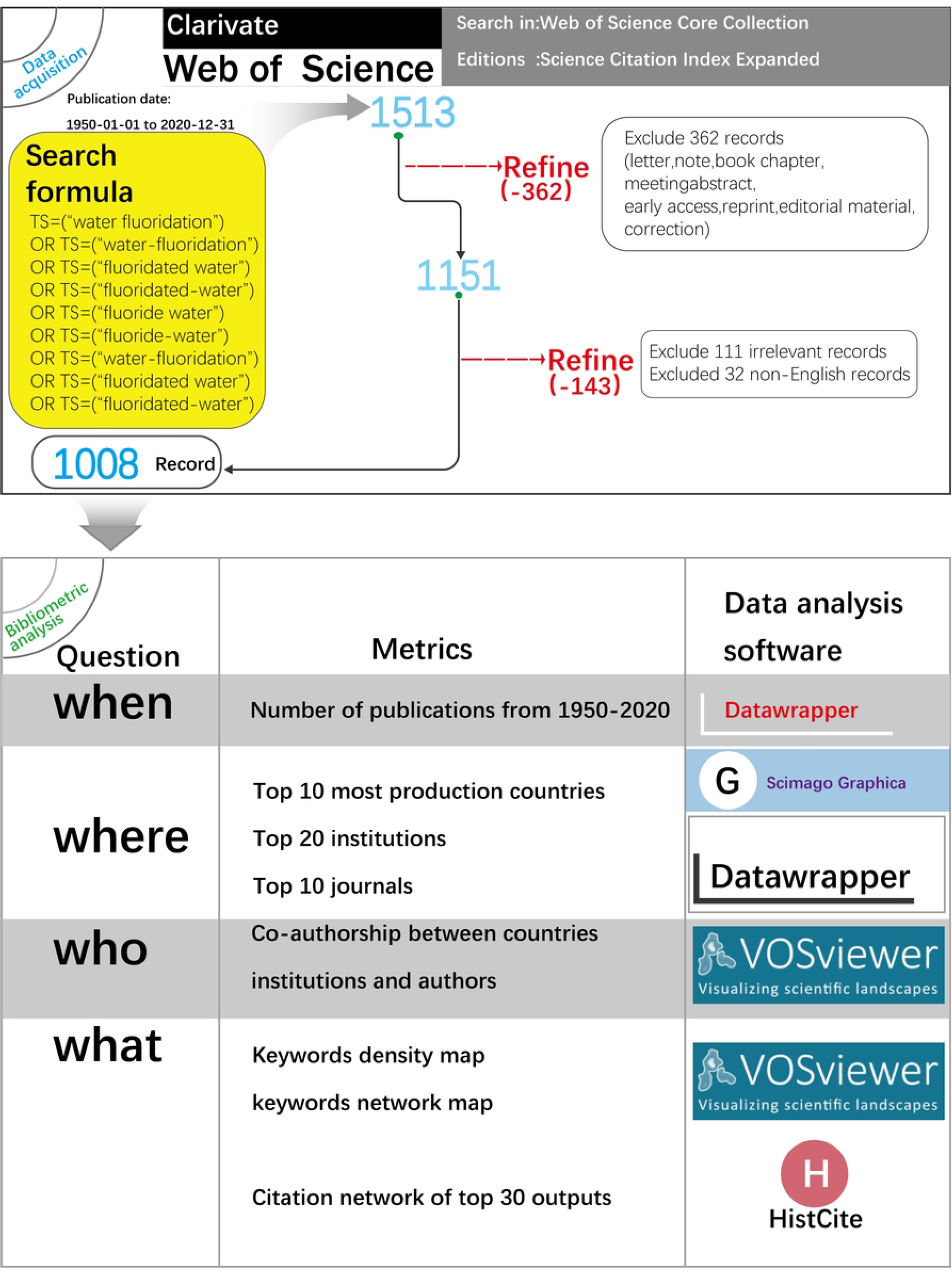
A flowchart representing retrieval strategy and structure diagram of this article.

### Top 10 most productive countries, 20 institutions, and 10 journals based on publication analyses

Through HistCite, we analysed the time of article publication, the 10 countries and 20 institutions with the largest number of articles, and the 10 journals with the largest number of WF articles. The impact factor (IF) and ranking of the journal are from the 2019 JCR report.

### Co-authorship between analysis and keyword analysis

In the cooperation network and keyword analyses, the size of the dot represents the frequency of the record; the higher the frequency, the larger dot. The connection between points is expressed by the thickness of the line; the more connections, the thicker the line. When we use VOSviewer for visualization, we merge synonyms. In keyword analysis, it is mainly the singular and plural forms of words or whether there are different forms caused by the “-” symbol between two words. In the author network, we merged different spelling forms of the same author and obtained the author’s unit information through the article (S1 Table).

### Analyses of the citation network of the top 30 most cited outputs

HistCite was used to analyse the citation relationships of the literature in the entire study, and the 30 documents with the highest total local citation scores (TLCSs) were incorporated into the citation chronology. The size of the point is related to the number of citations of this document in the research; the greater the number of points there are, the larger the outline of the point. A line indicates that there is a citation relationship between two documents. The arrow points from the citing document to the cited document. TLCS and total global citation score (TGCS) are the two most important parameters in HistCite. The TLCS is the number of times an article has been cited in the current database. The TGCS is the total number of citations in the WoS database. The TLCS can better reflect the influence of the article in terms of local research.

## Results

### Numbers of publication and the top 10 most production countries

From 1950 to 2020, 2,646 authors published a total of 1,008 articles in 294 journals. From 1950 to 1989, the average annual number of articles published did not exceed 4, but since 1990, the number of articles issued has increased significantly. From 1990 to 1999, 18.1 articles were published on average each year; from 2000 to 2009, 27.1 articles were published on average each year; and from 2010-2020, 38.9 articles were published on average each year (Figure 2a). The top 10 countries with the largest number of articles published a total of 810 articles, accounting for 80% of the total. Among them, the United States ranked first, with 269 articles. Australia ranked second with 143 articles, and the United Kingdom ranked third with 109 articles (Figure 2b).

**Figure 2.**
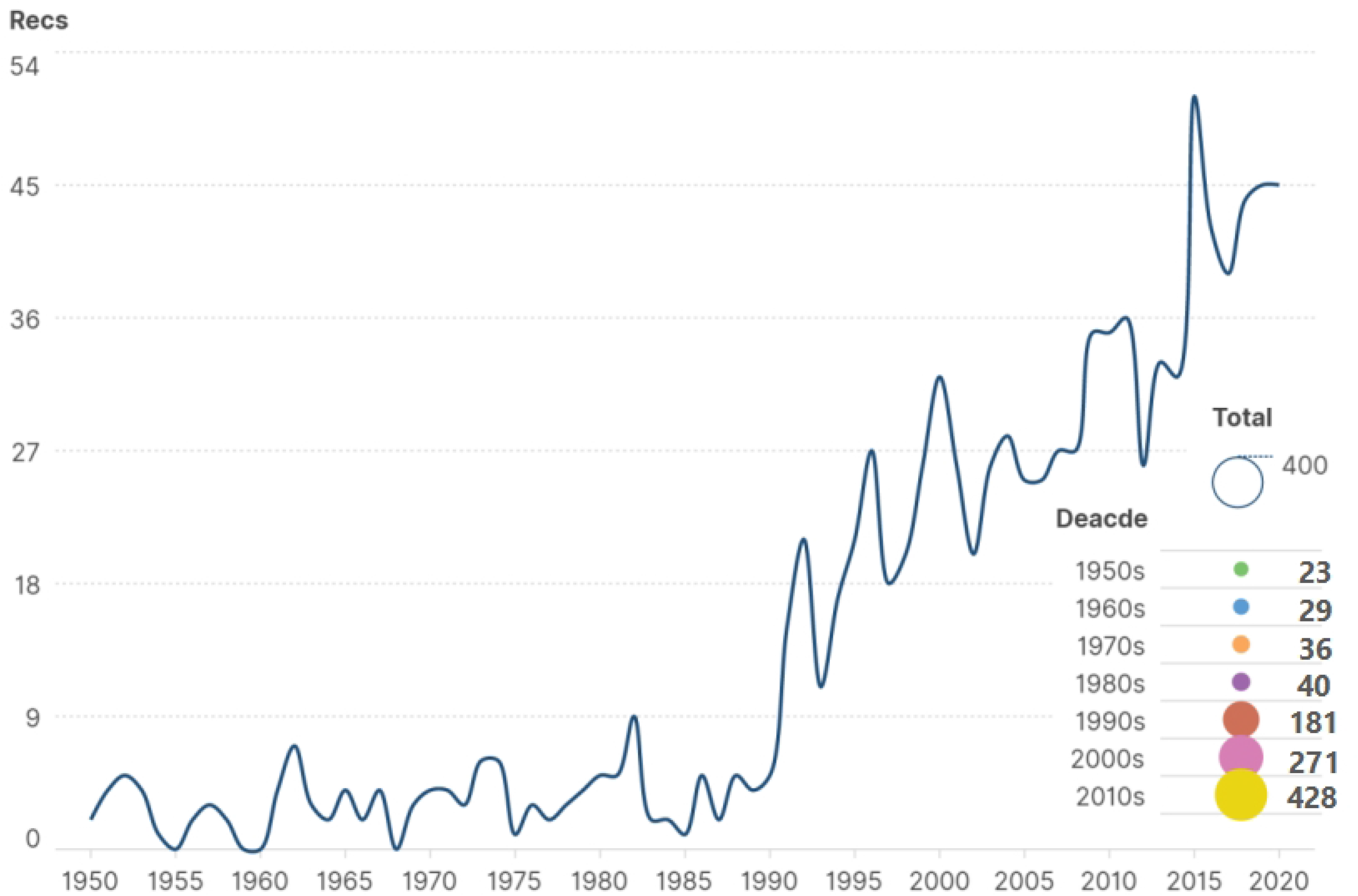
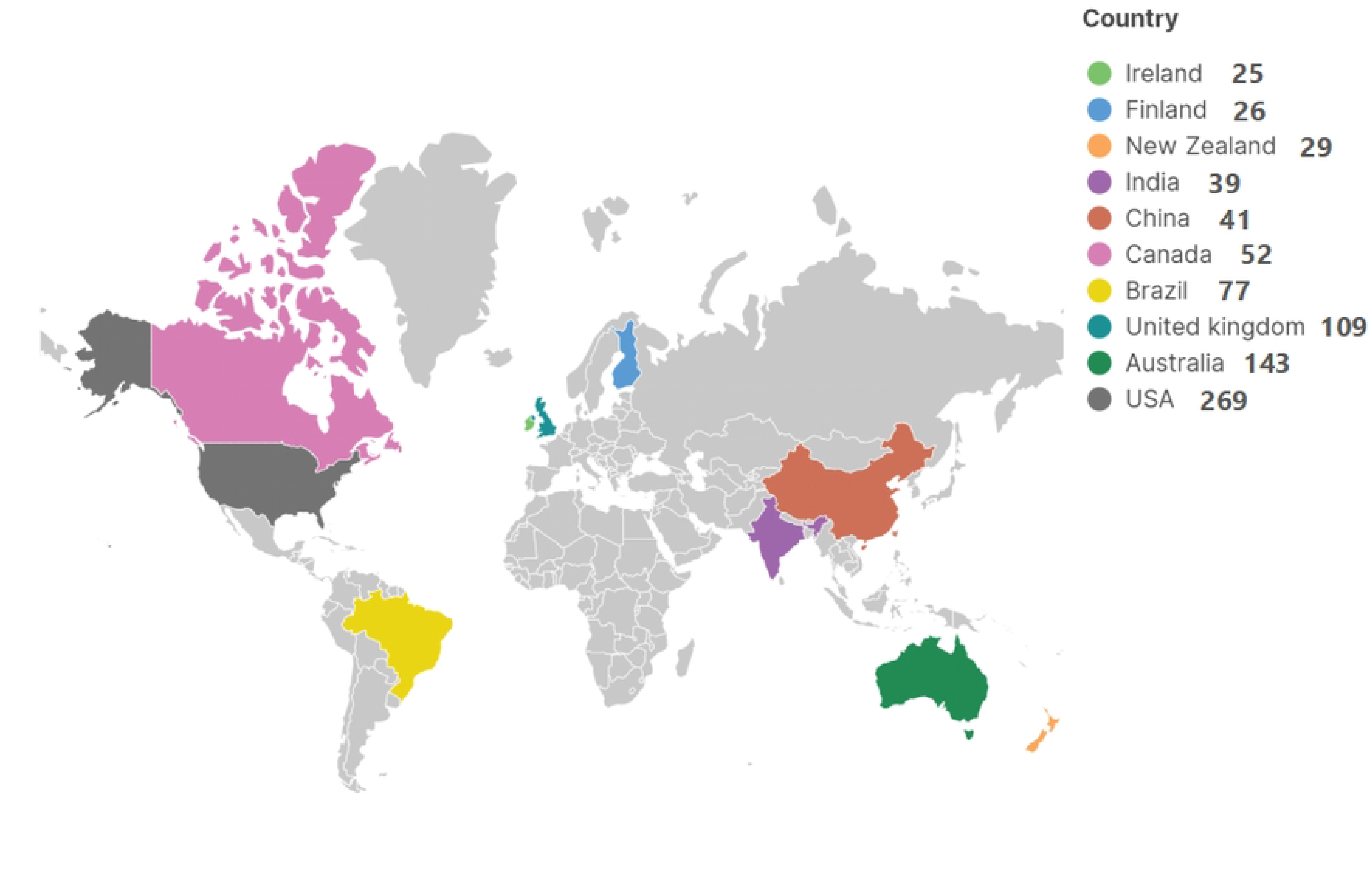
Number of publication and the top 10 most production countries. **A. The number of publications from 1950 to 2020.** **B. Top 10 most production countries.**

### Top 20 institutions ranked by the number of publications

In total, 335 articles were published among the top 20 institutions. Seven of these institutions are in the United States, and six are in Australia. Brazil, the United Kingdom, and Canada each have two institutions, and South Korea has one institution in the top 20. The number one publication institution is Australia’s University of Adelaide, with a total of 50 articles. The second is Brazil’s University of Sao Paulo, with 30 articles; and the third is Australia’s University of Melbourne, with 26 articles. Among them, the TLCS of the University of Adelaide is 218, the TGCS is 784 and is ranked first, and the TLCS per paper is 4.36, which means that the average article ranks fourth in terms of influence in this field. Moreover, we noticed that the U.S. CDC ranked first in terms of the influence of each article, with a TLCS per paper of 6.13 and a TGCS per paper of 29.19. The TLCS per paper indicates the influence of a single article in this field; the second and third places are the University of Iowa (United States) with a TLCS per paper of 5.73 and the University of British Columbia (Canada) with a TLCS per paper of 5.73, respectively (Figure 3).

**Figure 3.**
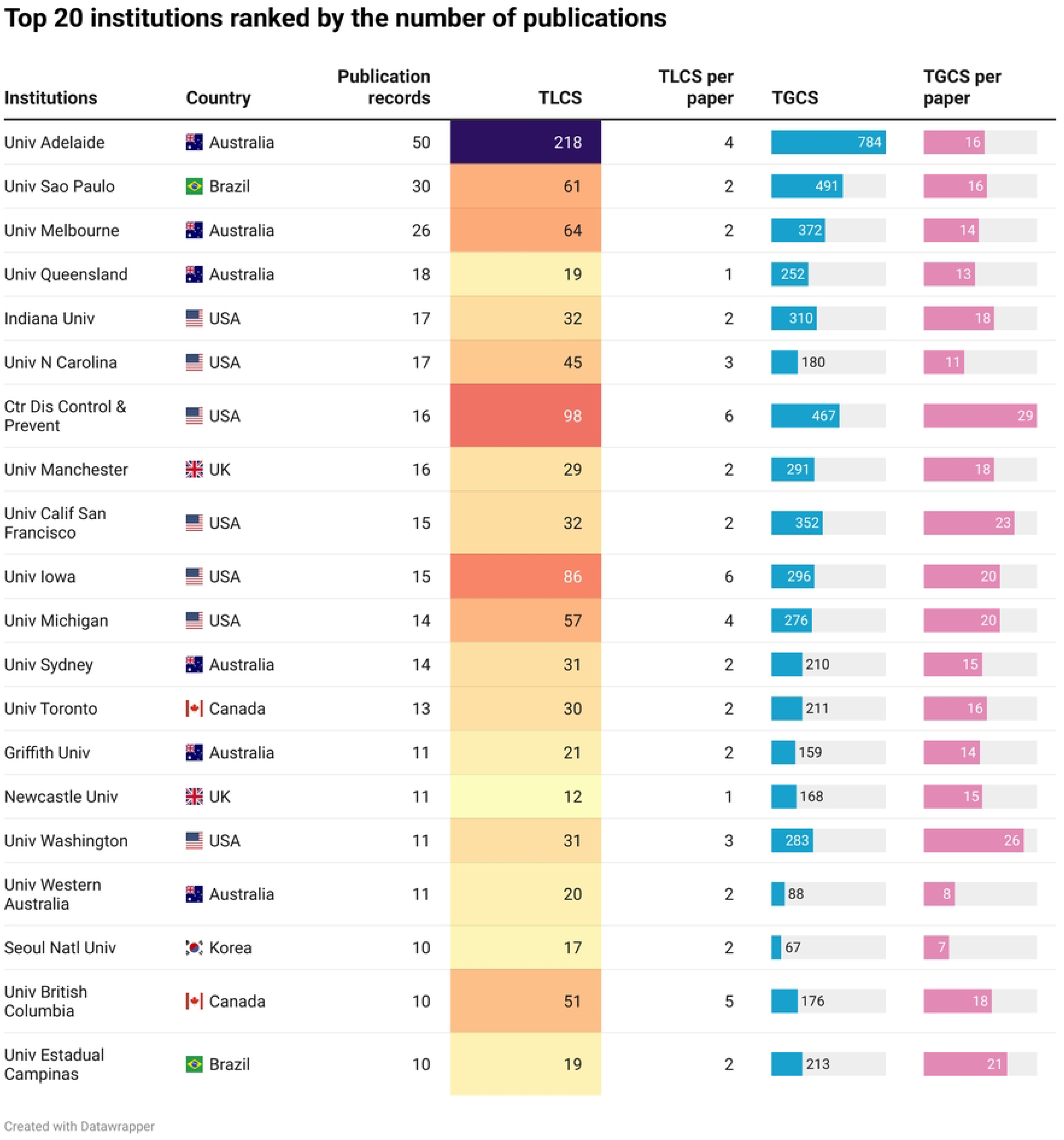
Top 20 institutions ranked by the number of publication.

### Top 10 journals ranked by number of publications

The top 10 journals published a total of 486 papers, accounting for 48% of the total number of articles. The total number of publications averages 48.6 articles per journal. The journals are classified on WoS mainly in Dentistry, Oral Surgery & Medicine and Public, Environmental & Occupational Health. *Community Dentistry and Oral Epidemiology* (2019 IF of 2.135) published 107 articles, ranking first. *Fluoride* and the *Journal of Public Health Dentistry* each published 75 related articles and tie for second place. According to the 2019 JCR report, among the 10 journals, the highest IF is 4.914 for the *Journal of Dental Research*, and the lowest is 0.679 (*Community Dental Health*). The average IF is 2.02. The TLCS of *Community Dentistry and Oral Epidemiology* is 472, and the TGCS is 1877; thus, both are ranked first among journals. However, the TLCS per paper for the *Journal of Dental Research* is 5.85, that for the *Journal of Public Health Dentistry* is 5.76, and that for *Community Dentistry and Oral Epidemiology* is 4.41, to round out the top three. The TGCS per paper of the *Journal of Dental Research* is 20.51, the TGCS per paper of *Community Dentistry and Oral Epidemiology* is 17.54, and the TGCS per paper of the *Caries Research* is 16.91; these journals are the top 3 in terms of TGCS per paper (Figure 4).

**Figure 4.**
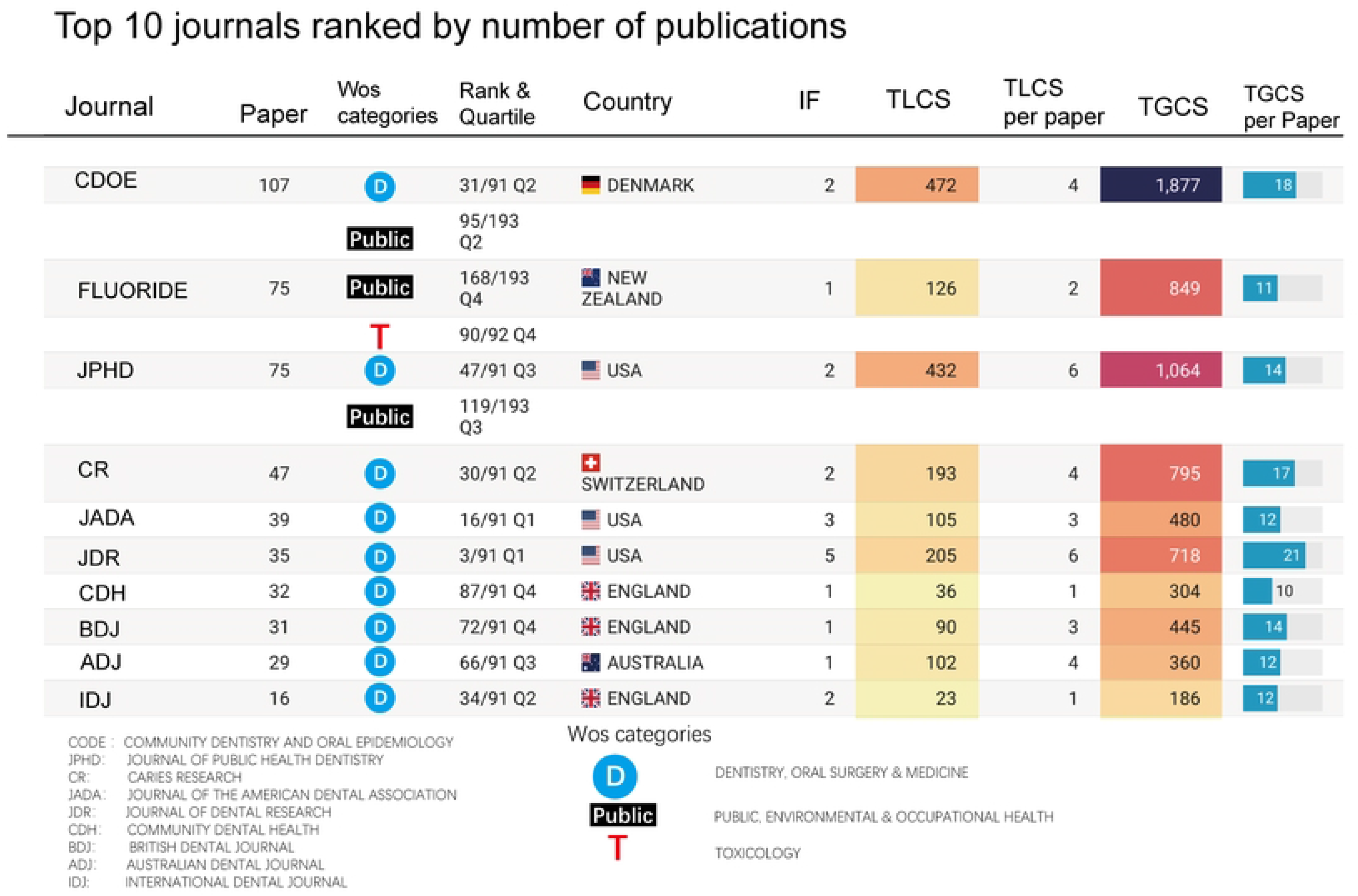
Top 10 journals ranked by number of publications.

### Co-authorship between countries, institutions, and authors

We can see that the United States has strong cooperative relations with Canada and Australia. The UK has strong cooperative relations with Ireland and Australia. Brazil has cooperative relationships with the United States and Australia, and its cooperative relationship with the United States is stronger than that with Australia (Figure 5a).

**Figure 5.**
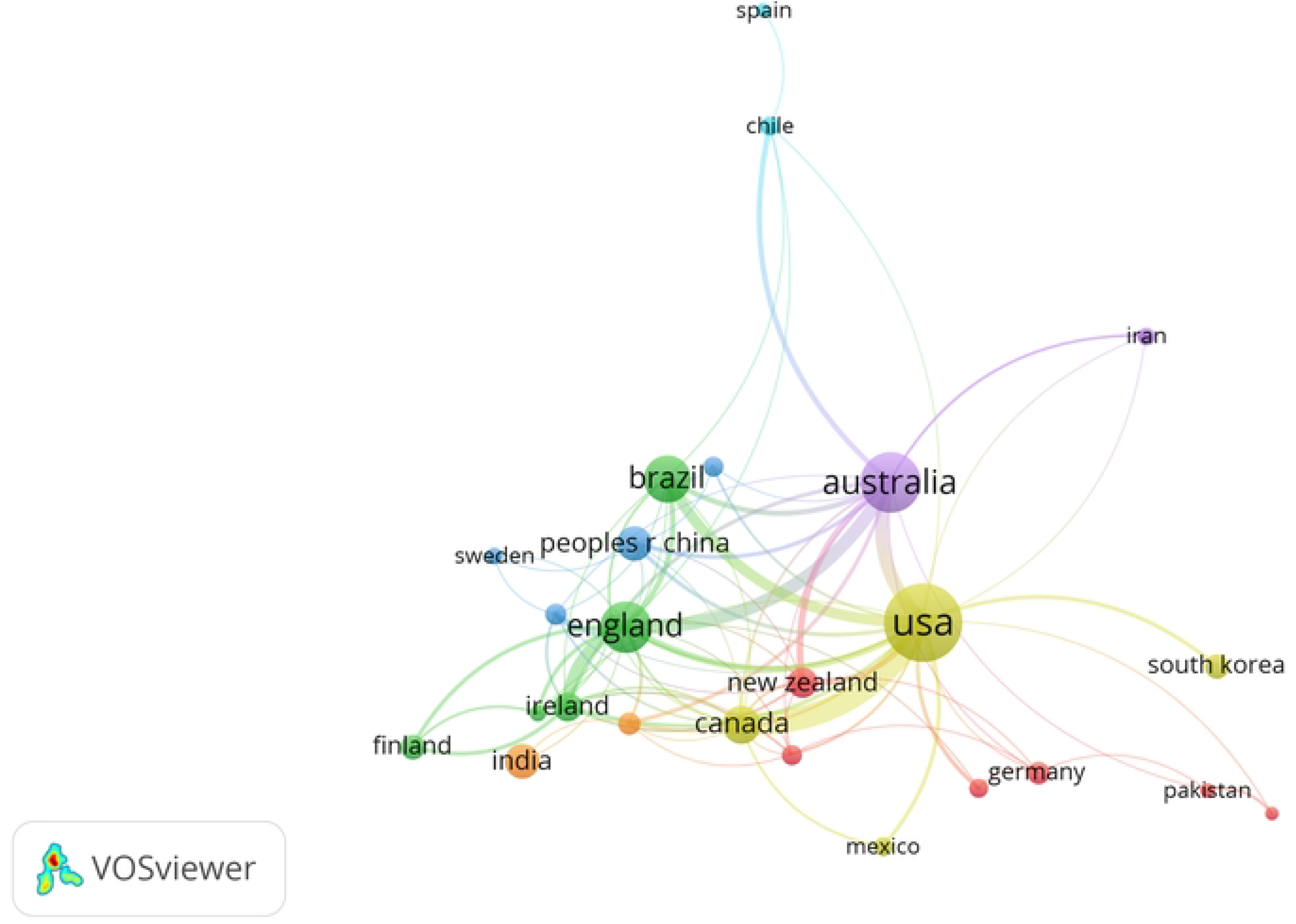
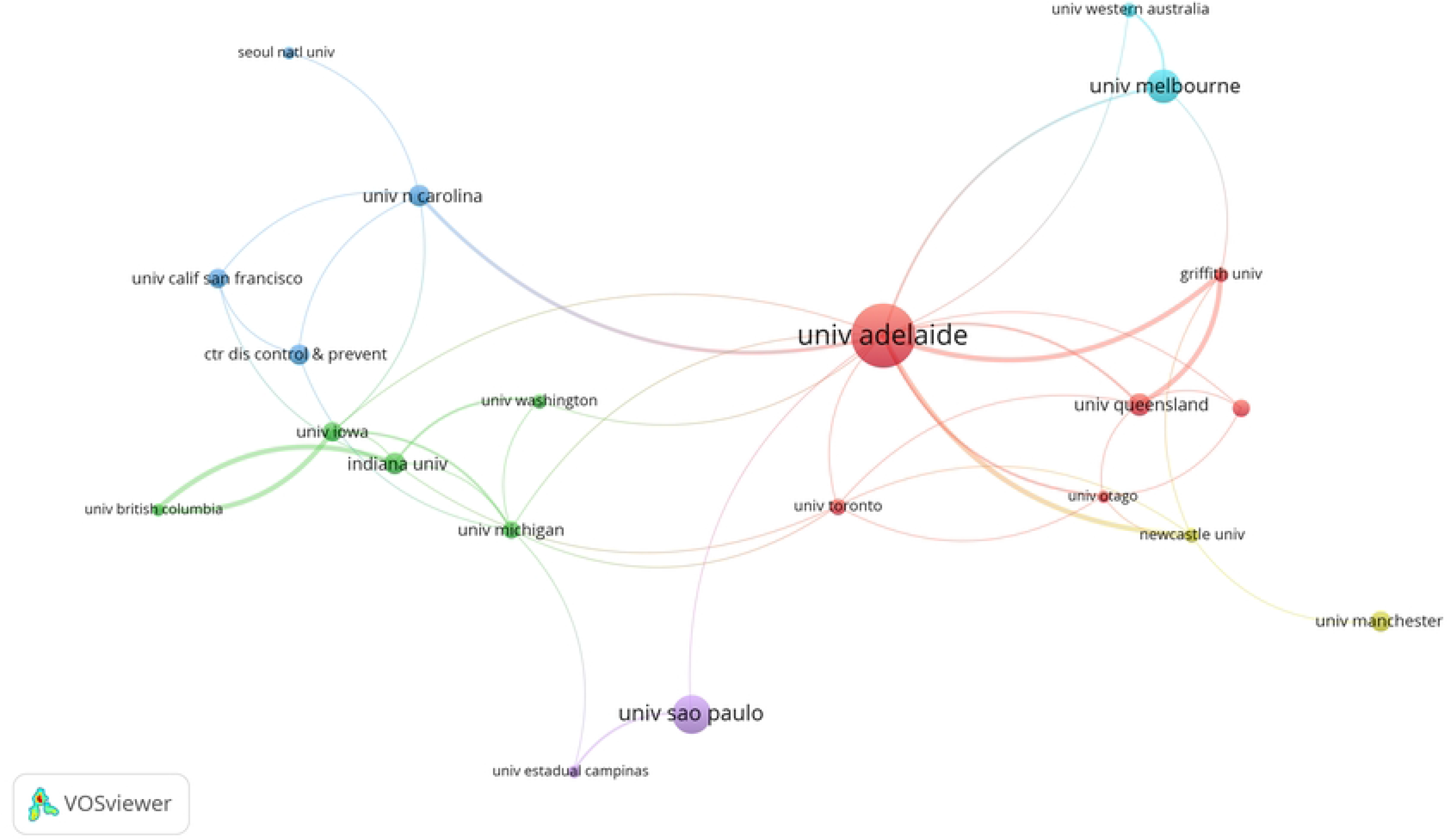
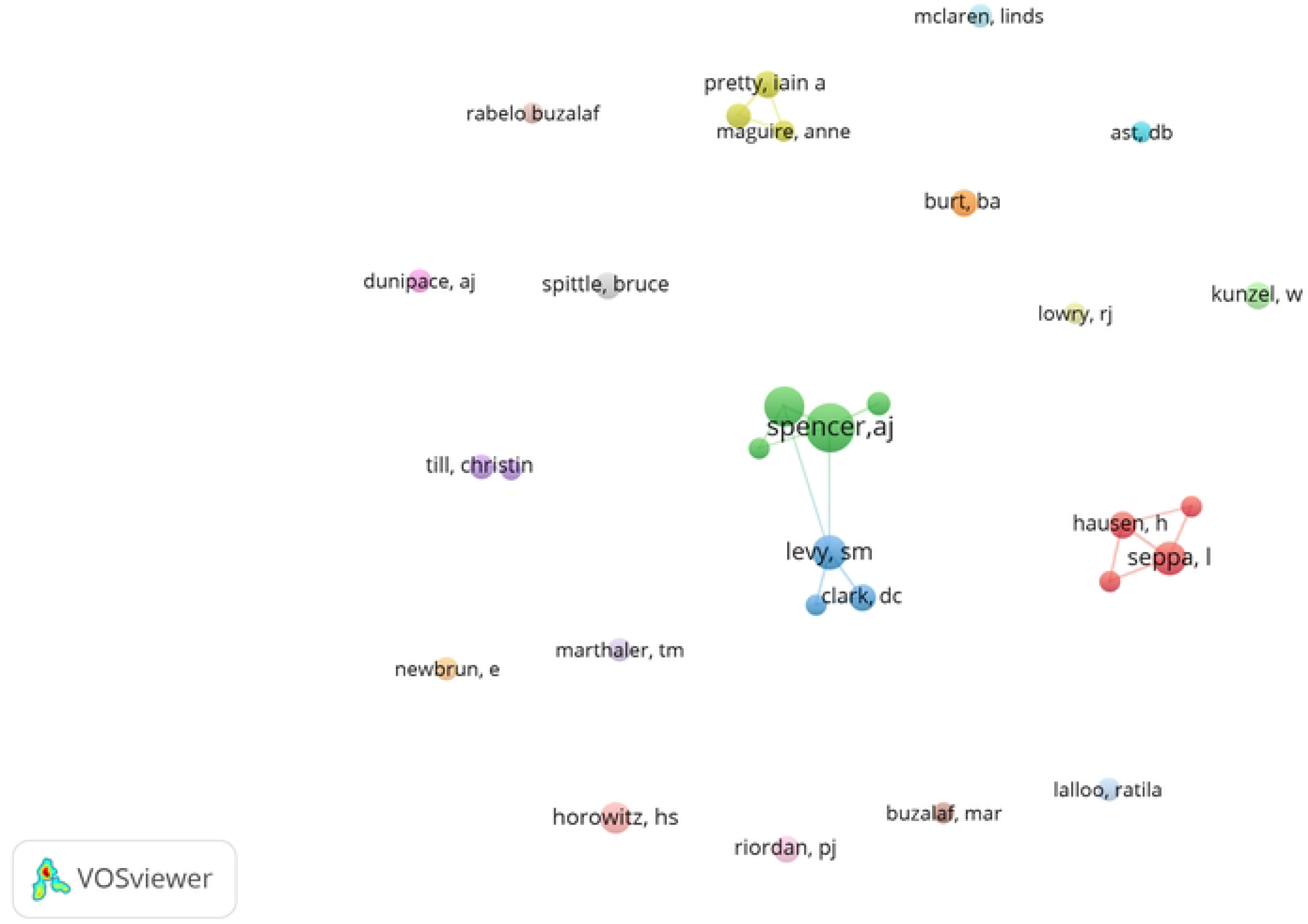
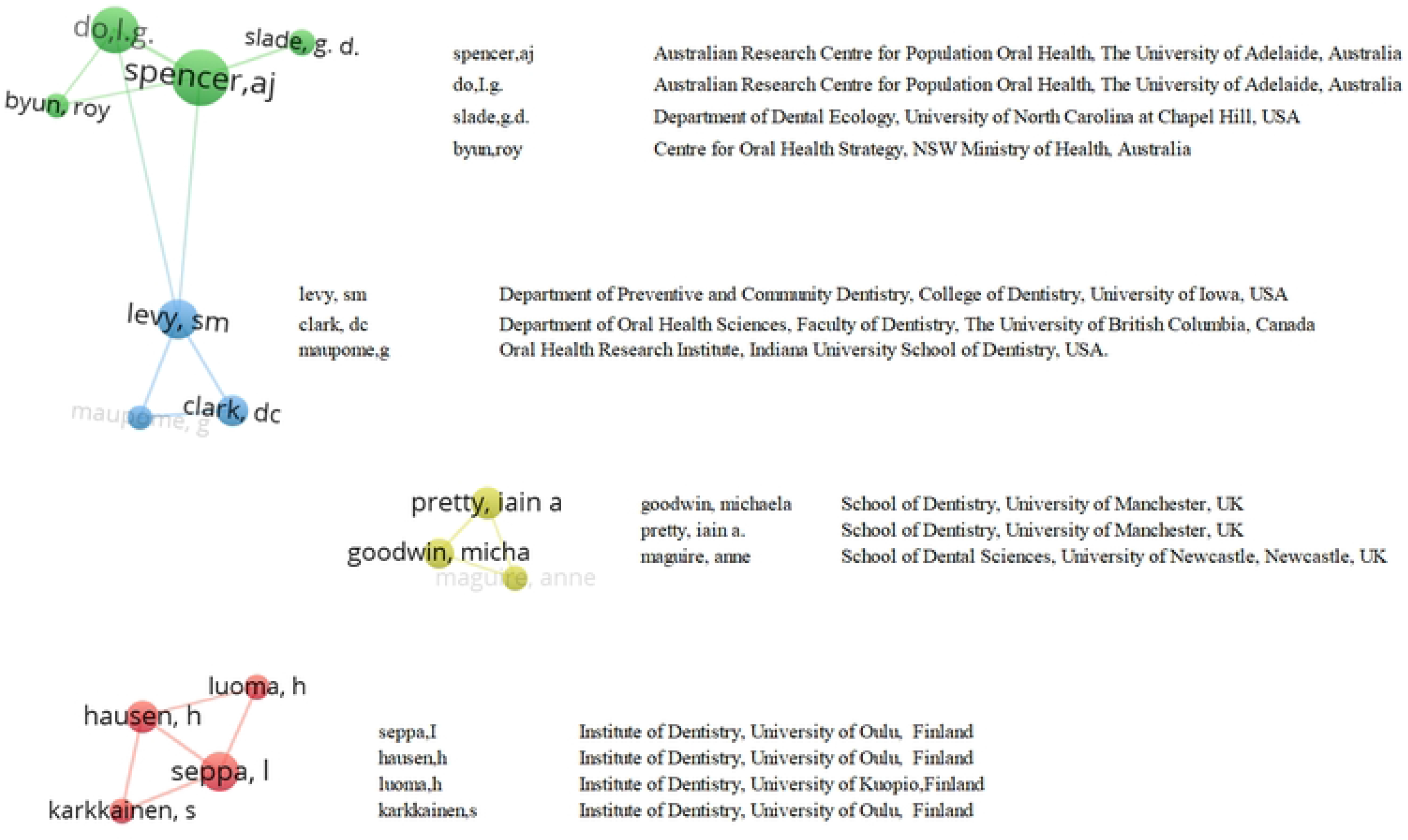
Co-authorship between countries(a), institutions(b), and authors(c,d) **A. Co-authorship between countries.** **B. Co-authorship between institutions.** **C. Co-authorship between authors.** **D. Detail of co-authorship network of more than 3 authors.**

It can be seen from the cooperation map of scientific research institutions that Australia’s Adelaide University is at the centre of the network. It has a strong cooperative relationship with the University of North Carolina, Griffith University, and Newcastle University. Moreover, it can be seen that the University of Queensland and Griffith University have a certain cooperative relationship. The University of British Columbia has a certain cooperative relationship with the University of Iowa and Indiana University (Figure 5b).

Co-authorship between authors is carried out by authors whose publication record is greater than or equal to 5, and there are only three co-authorship networks with more than three authors (Figure 5c). Seven scholars, including AJ Spencer and I Do from Adelaide University (Australia), SM Levy from Iowa University (United States) and DC Clark from the University of British Columbia (Canada), constitute the largest author collaboration network.

Seppa, Hausen, and Karkkainen from the University of Oulu (Finland) and Iuoma from the University of Kuopio (Finland) constitute the second largest cooperative network. Moreover, we noticed that Seppa and Hausen also been published research papers with Iuoma with the University of Kuopio.^13,14^

The last collaborative network is composed of three British scholars: Goodwin from the University of Manchester and Michaela and Maguire from the University of Newcastle. Figure 5d is the detail information of co-authorship network of more than 3 authors.

### Keyword analysis

Through the keyword density map, we can see that the main keywords are dental caries, fluoride, children, drinking water, prevalence, and dental fluorosis (Figure 6a).

**Figure 6.**
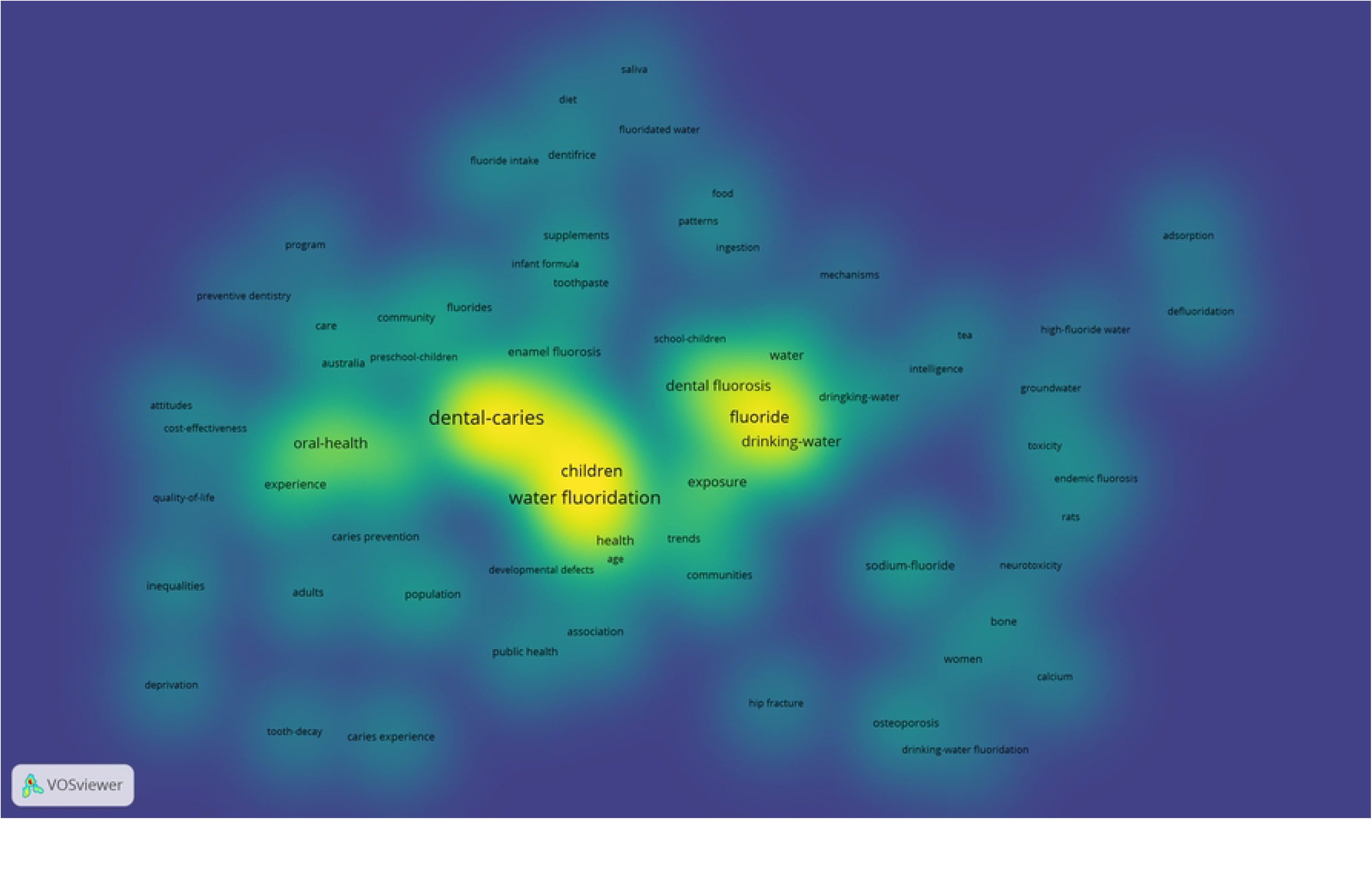
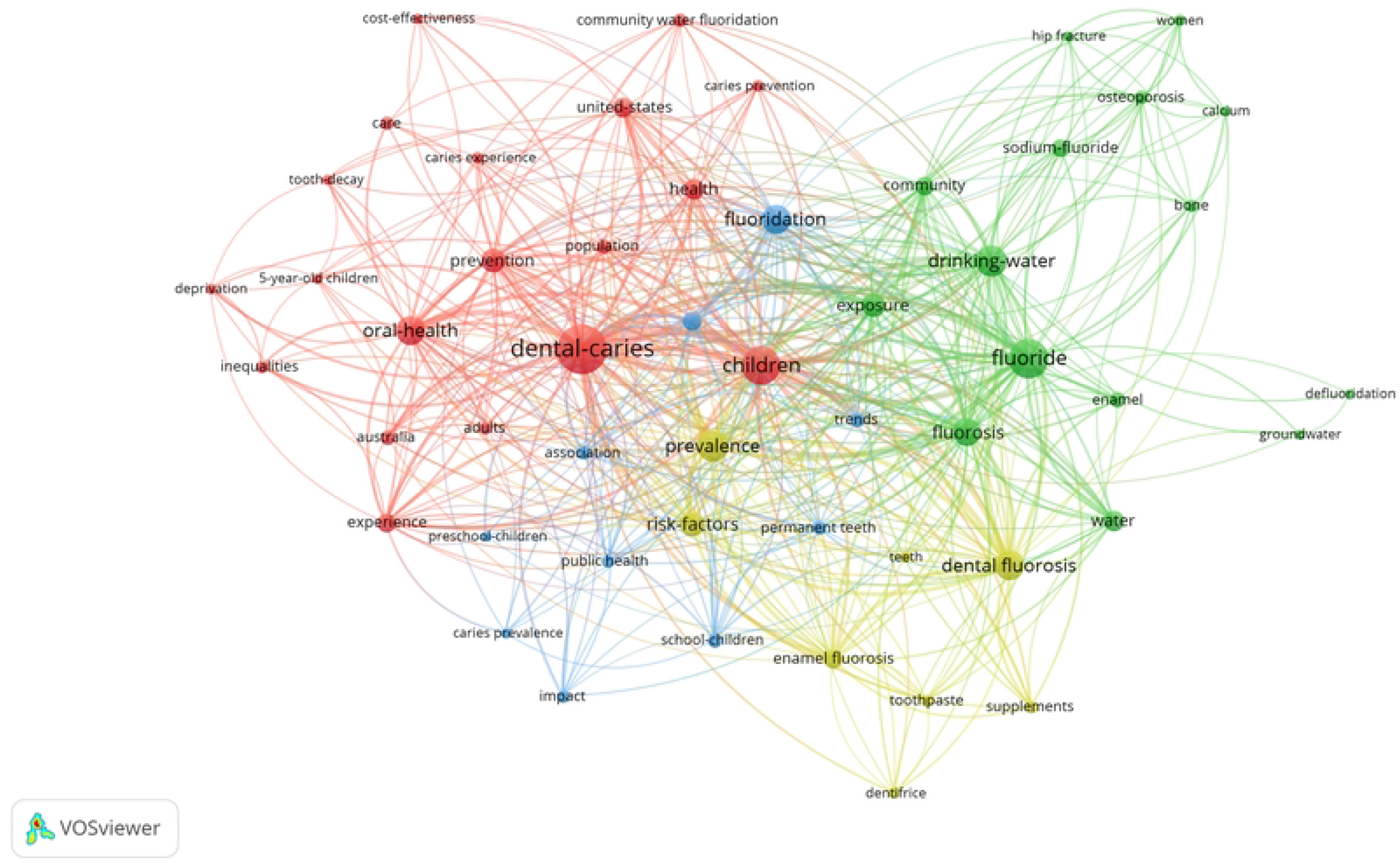
Keyword analysis. **A. Keywords density map.** **B. Keywords network map.**

In the keyword network diagram, VOSviewer divides the keywords into 4 clusters. Cluster 1 is red, and the high-frequency keywords are dental caries (321 times), children (187 times), oral health (112 times), and prevention (75 times), which are mainly related to dental caries. Cluster 2 is green. The high-frequency words were fluoride (203 times), drinking water (125 times), fluorosis (105 times), and exposure (72 times), which were mainly related to fluoride and fluorosis. Cluster 3 is blue, among which the high-frequency words are fluoridation (110 times) and epidemiology (46 times), which are mainly related to epidemics. Cluster 4 is yellow. The high-frequency keywords are prevalence (125 times), dental fluorosis (119 times), risk factors (79 times), and enamel fluorosis (45 times), which are mainly related to dental fluorosis (Figure 6b).

### Citation network of top 30 most cited outputs

In total, 30 records were selected for citation analysis (Figure 7). The minimum TLCS in this group was 16, and the maximum TLCS was 82.

**Figure 7.**
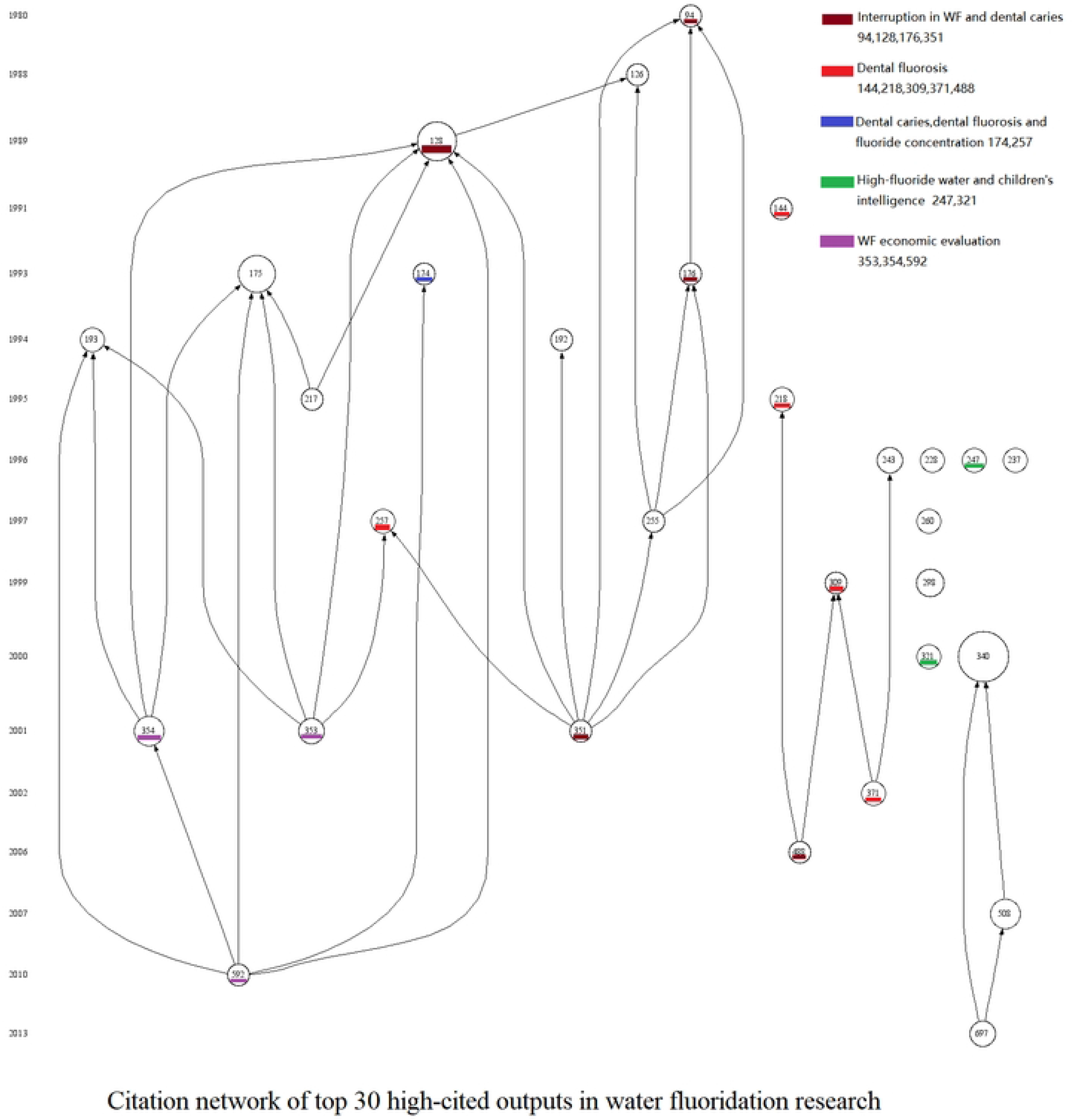
Citation network of top 30 high-cited outputs in WF.

The first highly cited article is from 1980, and the last one is from 2013. The articles were mainly published in the Journal of Public Health Dentistry (8 articles) and Community Dentistry and Oral Epidemiology (6 articles). The highest TLCS is achieved by Systematic Review of Water Fluoridation (record 340) published in the British Medical Journal in 2000.^15^ Records 144, 218, 309, 371, and 488 are all focused on dental fluorosis, of which 218 and 488 studied enamel fluorosis on the maxillary incisors.^16,17^ Records 94, 128, 176, and 351 studied the relationship between interrupted WF and dental caries. Records 174 and 257 studied the relationship between dental caries, dental fluorosis and fluoride concentration. Records 247 and 321 studied the relationship between high-fluoride water and children’s intelligence. Records 354, 353, and 592 are all WF economic evaluations. In addition, we noticed that record 353 has been withdrawn.^18^ S2 Table shows the specific information of the top 30 highly cited outputs.

## Discussion

WF-related research has attracted the attention of top medical journals since 1950.^19-24^ Major institutions such as the U.S. Public Health Service,^25^ National Research Council,^26^ World Health Organization,^27^ and Institute of Medicine^28^ have confirmed the safety of WF, but in the past three years, they have continuously reported on WF investigations based on social media, showing that the public still has doubts regarding WF.^7,29^ Hyo-Jung Oh et al (2018) performed a trend analysis on studies in the literature related to WF and caries but did not analyse other related WF literature and did not have detailed indicators such as highly cited literature and cooperative relationship analysis.^30^

This article carried out bibliometric research and analysis on articles published in the field of WF over the past 70 years. After 1991, the number of articles published increased sharply. From 1991 to 2010, the average annual number of publications exceeded 20, and the number of publications in the last 10 years was 39.3. The United States and Australia are the most productive countries with the most productive scientific research institutions. Australia’s Adelaide University ranks first with 50 papers and ranks first in the world in terms of the TLCS and TGCS and fourth in the world in terms of the TLCS per paper. This finding shows that Adelaide University not only publishes a large number of articles but also publishes high-quality papers and has an important influence in this field. The main co-authorship network between organizations also confirms this point. The United States and Australia have more co-authorships. Adelaide University is at the centre of the institutional cooperation network. At the same time, AJ Spencer and IG GO (Adelaide University), SM Levy (Iowa University, USA), and DC Clark (University of British Columbia, Canada) constitute the largest author network in the co-authorship analysis.

According to the WoS classifications, WF research is mainly published in Dentistry, Oral Surgery & Medicine. *Community Dentistry and Oral Epidemiology* published 107 articles, ranked first in the number of articles published and in the TLCS and TGCS. In addition, ranked thrid in the TLCS per paper index. In summary, it can be seen that *Community Dentistry and Oral Epidemiology* has an important position in this research field. However, in terms of the impact of a single article in this research field, the *Journal of Dental Research* ranks first in the top 10 journals, showing its important influence in the field of dentistry.

Analysing the keyword network in the WF literature, the results show that the most common keywords are dental caries, fluoride, children, drinking water, prevalence, and fluorosis. The keywords of WF mainly include prevention of dental caries, systemic toxicity of fluoride, dental fluorosis and epidemiological investigation. The chronological chart of the top 30 highly cited articles in the field shows that the most cited articles are mainly focused on 1) dental fluorosis, especially fluorosis of the maxillary central incisor; 2) the relationship between interruption WF and caries; 3) the impact of drinking water on children’s intelligence; and 4) economic evaluation.

This is the first article to report on the bibliometrics of WF. The list of highly cited articles will provide an important source of information for researchers, public health personnel and government agencies. Analysis of research hotspots in this field, publications in journals, analysis of high-yield institutions and cooperation relationships will be beneficial to later scientific research and cooperation choices of researchers and government agencies.

Some limitations still exist in our studies. The data in this paper only comes from Wos, and there is no search on other databases such as Pubmed. At the same time, the deadline for data inclusion in this paper is 2020, and the lack of research data in the past three years will have a certain impact on the data in this paper.However, the WoS database is considered the best database for this type of evaluation because the use of additional biomedical databases does not significantly increase the production of related journals.^31^ Secondly, we are exploring the 70 years of research changes from 1950 to 2020, in which the chronological statistics are measured in 10 years. Hence, there is no further addition of articles from the last 3 years.

## Data Availability

All relevant data are within the manuscript and its Supporting Information files.

## Acknowledgements

The authors would like to acknowledge the general support of Xuzhou Medical University and Chongqing Medical University.

## Supporting information

**S1 Table. Synonym merging.**

**S2 Table. Specific information of highly cited 30 outputs.**

## References

1. Over 75 years of Community Water Fluoridation, https://www.cdc.gov/fluoridation/basics/anniversary.htm (archived on 18 Mar 2021).

2. Healthy People 2020: health communication and health information technology, https://www.healthypeople.gov/2020/topics-objectives/topic/oral-health/ objectives (archived on 26 Aug 2019).

3. Centers for Disease Control and Prevention (CDC). Ten great public health achievements--United States, 1900-1999. MMWR Morb Mortal Wkly Rep. 1999;48(12):241-3.

4. Mackert M, Bouchacourt L, Lazard A, Wilcox GB, Kemp D, Kahlor LA, et al. Social media conversations about community water fluoridation: formative research to guide health communication. J Public Health Dent. 2021;81(2):162–166.

5. Schluter PJ, Hobbs M, Atkins H, Mattingley B, Lee M. Association Between Community Water Fluoridation and Severe Dental Caries Experience in 4-Year-Old New Zealand Children. JAMA Pediatr. 2020;174(10):969–976.

6. Morabia A. Community Water Fluoridation: Open Discussions Strengthen Public Health. Am J Public Health. 2016;106(2):209–10.

7. Oh HJ, Kim CH, Jeon JG. Public Sense of Water Fluoridation as Reflected on Twitter 2009-2017. J Dent Res. 2020;99(1):11–17.

8. Zhu R, Wang Y, Wu R, Meng X, Han S, Duan Z. Trends in high-impact papers in nursing research published from 2008 to 2018: A web of science-based bibliometric analysis. J Nurs Manag. 2020;28(5):1041–1052.

9. Curt C. Multirisk: What trends in recent works? - A bibliometric analysis. Sci Total Environ. 2021;763:142951.

10. Glanville J, Kendrick T, McNally R, Campbell J, Hobbs FD. Research output on primary care in Australia, Canada, Germany, the Netherlands, the United Kingdom, and the United States: bibliometric analysis. BMJ. 2011;342:d1028.

11. van Eck NJ, Waltman L. Software survey: VOSviewer, a computer program for bibliometric mapping. Scientometrics. 2010;84(2):523–538.

12. Garfield E. From the science of science to Scientometrics visualizing the history of science with HistCite software. J Informetr 2009;3(3):173–179.

13. Seppä L, Luoma H, Hausen H. Fluoride content in enamel after repeated applications of fluoride varnishes in a community with fluoridated water. Caries Res. 1982;16(1):7–11.

14. Seppä L, Hausen H, Luoma H. Relationship between caries and fluoride uptake by enamel from two fluoride varnishes in a community with fluoridated water. Caries Res. 1982;16(5):404–12.

15. McDonagh MS, Whiting PF, Wilson PM, Sutton AJ, Chestnutt I, Cooper J, et al. Systematic review of water fluoridation. BMJ. 2000;321(7265):855-9.

16. Evans RW, Darvell BW. Refining the estimate of the critical period for susceptibility to enamel fluorosis in human maxillary central incisors. J Public Health Dent. 1995;55(4):238–49.

17. Hong L, Levy SM, Broffitt B, Warren JJ, Kanellis MJ, Wefel JS, et al. Timing of fluoride intake in relation to development of fluorosis on maxillary central incisors. Community Dent Oral Epidemiol. 2006;34(4):299–309.

18. Griffin SO, Gooch BF, Lockwood SA, Tomar SL. Quantifying the diffused benefit from water fluoridation in the United States. Community Dent Oral Epidemiol. 2001;29(2):120–9.

19. KNUTSON JW. The case for water fluoridation. N Engl J Med. 1952;246(19):737–43.

20. Cooper C, Wickham CA, Barker DJ, Jacobsen SJ. Water fluoridation and hip fracture. JAMA. 1991;266(4):513–4.

21. Danielson C, Lyon JL, Egger M, Goodenough GK. Hip fractures and fluoridation in Utah’s elderly population. JAMA. 1992;268(6):746–8.

22. Jacobsen SJ, O’Fallon WM, Melton LJ 3rd. Hip fracture incidence before and after the fluoridation of the public water supply, Rochester, Minnesota. Am J Public Health. 1993;83(5):743–5.

23. Gibson SL, Gibson RG. Water fluoridation. Clearer evidence of benefits and risks is needed. BMJ. 2001;322(7300):1487.

24. Yeung CA. Water fluoridation could save NHS millions every year. BMJ. 2014;348:g2855.

25. Public Health Service report on fluoride benefits and risks. MMWR Recomm Rep. 1991;40(RR-7):1-8.

26. National Research Council Committee on Toxicology. Health effects of ingested fluoride. Washington, DC: National Academy Press, 1993.

27. World Health Organization. Environmental health criteria 36: fluorine and fluorides. Geneva: World Health Organization, 1984.

28. Institute of Medicine (US) Standing Committee on the Scientific Evaluation of Dietary Reference Intakes. Dietary Reference Intakes for Calcium, Phosphorus, Magnesium, Vitamin D, and Fluoride. Washington (DC): National Academies Press (US); 1997.

29. Mackert M, Bouchacourt L, Lazard A, Wilcox GB, Kemp D, Kahlor LA, et al. Social media conversations about community water fluoridation: formative research to guide health communication. J Public Health Dent. 2021;81(2):162–166.

30. Oh HJ, Choi HM, Kim C, Jeon JG. Trend Analysis of Studies on Water Fluoridation Related to Dental Caries in PubMed. Caries Res. 2018;52(6):439–446.

31. Aggarwal A, Lewison G, Idir S, Peters M, Aldige C, Boerckel W, et al. The state of lung cancer research: a global analysis. Journal of Thoracic Oncology. 2016;11:1040–50.

